# Development and validation of a productive liquid chromatography-tandem mass spectrometry method for the analysis of androgens, estrogens, glucocorticoids and progestagens in human serum

**DOI:** 10.1101/2021.04.12.21255305

**Authors:** Michele Iannone, Anna Pia Dima, Francesca Sciarra, Francesco Botrè, Andrea M. Isidori

## Abstract

Adrenal and gonadal disorders are very often coupled, due to common etiology or pathophysiology. We present the development, validation and application of a liquid chromatography tandem mass spectrometry (LC-MS/MS) method for the simultaneous analysis of androgens (androstenedione (A4), testosterone (T), dihydrotestosterone (DHT), and dehydroepiandrosterone sulfate (DHEA-S)), estrogens (estrone (E1), estradiol (E2), estriol (E3)), glucocorticoids (cortisol (F), cortisone (E), corticosterone (B), 11-deoxycortisol (S), 21-deoxycortisol (21DF), 11-deoxycorticosterone (11DB)), and progestagens (progesterone (P4), 17α-hydroxyprogesterone (17OHP4) and 17α-hydroxypregnenolone (17OHP5)) in human serum for clinical use. Samples (250 &[mu]L of matrix) spiked with isotopic labelled internal standards were extracted with tert-butylmethyl ether (TBME) prior to LC-MS/MS analysis. The chromatographic separation of the underivatized endogenous steroids was achieved on a reversed-phase column (C18 Zorbax Eclipse Plus) using a methanol-water gradient. The LC column was coupled to a triple quadrupole mass spectrometer equipped with an electrospray (ESI) source operating both in positive and in negative mode, with acquisition in multiple reaction mode. The method was validated using surrogated matrices and human serum samples. The proposed method was proven to be specific for all the considered steroids; and linearity was also assessed (R^2^ > 0.99) in the ranges of quantification investigated. The lower limits of quantification (LLOQs) were in the range of 10 - 400 pg/mL depending on the target steroid. Accuracy was in the range 80 - 120% for all the target compounds, the extraction recovery was higher than 65% for all the steroids considered and no remarkable matrix effect, expressed in terms of ion enhancement and ion suppression, was observed. To test the reliability of the developed and validated method, the analysis of serum samples collected from ten healthy subjects (5M/5F) was performed. In the clinical settings there is a growing need to develop accessible methods for full steroid hormone profiling. The dynamic link between steroidogenic glands and liver enzymatic processing (activation and clearance) attributes to the profile a much greater clinical meaning than a set of individually measured hormones. The presented method can be used to identify trajectories of deviation from the concentration normality ranges applied to disorders of the gonadal and adrenal axes.

## Introduction

Steroid hormones are a class of endogenous substances characterized by a sterane skeleton composed by 4 fused rings (three cyclohexane and one cyclopentane), sustaining important role in various physiological functions (e.g., development, sexual differentiation, reproduction, pregnancy, stress, metabolism, immune modulation). The biosynthesis of steroid hormones occurs only in highly specialized tissues (i.e., adrenal cortex glands, gonads, placenta).Their biological effects, however, are significantly influenced by peripheral tissue enzymatic conversion. For example, cortisol can be inactivated and re-activated by the 11-beta-hydrosteroid-dehidrogenase type 1 in the liver (Isidori, 2003), while testosterone can be aromatized into estradiol or reduced to di-hydro-testosterone (Othonos, 2020), triggering important systemic effects. The precise and accurate quantification of endogenous steroid hormones, and their intermediate products, in plasma, serum or dried blood spots (DBSs) represents a powerful tool for the investigation of the hormone status, the early identification of endocrinological disorders, and follow-up under therapeutic interventions (Giannetta 2012; Pofi, 2018). For indeed, steroid diagnostic is also employed for the diagnosis of a series of adrenal (Minnetti, 2020), sexual (Aversa, 2004), and reproductive disorders (Isidori, 2006), including those related to sexual differentiation, gonadal function as well as precocious puberty and polycystic ovary disease (PCOS) (Minutti, 2004; Peitzsch, 2015; Handelsman, 2017; Taylor, 2017).

In clinical laboratories, the routine methods mainly used since now for the detection and quantitation of endogenous steroids in blood (serum and/or plasma) are immunoassays, which can utilize either radioactive or non-radioactive markers, and belong to the excess-antigen/limited-antibody type immunoassays. These analytical methods, which in most of the cases are available as kits, are used because of their noteworthy advantages in terms of costs and automation, but, at the same time, they present a series of major limitations (Auchus, 2014; Handelsman, 2017). Commercial diagnostic kits, in fact, are often characterized by poor antibody specificity, due to the cross reactivities with structurally similar metabolites (poor accuracy) especially in the low concentration ranges; furthermore, they present limitation in sensitivity, may be affected by matrix interferences (which cannot be corrected by the use of suitable internal standards); finally, their dynamic range is quite limited and highly variable among different kits and different laboratories. Another serious drawback of immunoassay-based methods is represented by the impossibility to perform multiple analytes analysis in the same session and on the same sample aliquot, leading to a significant waste of sample volume and longer overall analysis time (Ismail, 2002; Stanczyk, 2003; Rosner, 2007; Koal, 2012), introducing errors in the relative ratios between hormones and their metabolites

Because of the limitations in terms of sensitivity, accuracy, precision and specificity of the immunoassay-based analytical method for the identification and quantification of endogenous steroids in blood matrices, numerous methods based on gas chromatography (GC) or liquid chromatography (LC) coupled to mass spectrometry (MS) have been developed for clinical applications in the last years. Chromatographic-mass spectrometric methods present significant improvements with respect to immunoassays in terms of selectivity and sensitivity (Shackleton, 1990; Wolthers, 1999; Vogaser, 2007; Soldin, 2009; Krone, 2010; Shackleton, 2010; Grebe, 2011; Kushnir, 2011; Carvalho, 2012; Keevil, 2016; Shackleton, 2018).

The GC-MS^n^ analysis of the urinary steroid profile, as well as of the urinary excretion of synthetic steroids, is an analytical approach that remains unrivaled in forensic sciences (i. e. antidoping analysis, toxicological applications) (Narducci, 1990; Buiarelli, 2001; Peng, 2002; Frati, 2015; Palermo, 2016; Stoll, 2020 a; Stoll, 2020 b; Iannone, 2020). At the same time, GC-MS has also been proven very useful for the unequivocal identification of different well-characterized steroid metabolic disorders, like 21-hydroxylase deficiency and hypogonadism (Caulfield, 2002; Di Luigi, 2009; Di Luigi, 2012; Iannone, 2019; Storbeck, 2019). However, in the last 10-15 years, GC-MS^n^ analysis of urinary endogenous steroids for clinical application has been progressively superseded by LC-MS^n^ or LC coupled to high resolution mass spectrometer (i. e. orbitrap, time-of-flight), whose applications also included the analysis of blood matrices. For indeed, methods based on liquid chromatography coupled to mass spectrometry allow to bypass the derivatization step, necessary in GC-MS^n^ analysis to generate volatile and thermally stable compounds. Furthermore, analysis of the target steroid levels in blood could also reduce the utility of the analysis of the entire volume of urine excreted in 24 hours (Ceglarek, 2009; Fanelli, 2011; Keski-Rahkonen, 2011; Gaudl, 2016; Matysik, 2016; Ponzetto, 2016; Ponzetto, 2017; Taylor, 2017; Hakkinen, 2018; Liu, 2019), with a significant improvement both in the logistics of sample collection and analysis, and in the rapidity of the result turnaround. Nonetheless, GC-MS^n^ still remains unrivaled in the case highest chromatographic resolution is essential, for instance for the unambiguous identification of synthetic steroid isomers.

To date, as reported in the recommendations of different Endocrine Societies (Isidori, 2015; Isidori, 2020), high-quality and well-validate LC-MS/MS methods for the identification and quantification of endogenous steroids in blood (serum and/or plasma), are the methods of choice for use in the clinical laboratory, thanks to the simultaneous detection of multi-class hormones during a single injection. We are here proposing the development, validation and application of a simplified liquid-chromatography tandem mass spectrometry (LC-MS/MS) method for the analysis of 16 endogenous steroid hormones belonging to the classes of androgens (androstenedione (A4), testosterone (T), dihydrotestosterone (DHT), and dehydroepiandrosterone sulfate (DHEA-S)), estrogens (estrone (E1), estradiol (E2), estriol (E3)), glucocorticoids (cortisol (F), cortisone (E), corticosterone (B), 11-deoxycortisol (S), 21-deoxycortisol (21DF), 11-deoxycorticosterone (11DB)), and progestagens (progesterone (P4), 17α-hydroxyprogesterone (17OHP4) and 17α-hydroxypregnenolone (17OHP5)) with the aim to obtain a pathway-driven serum steroid profile which could be used in clinical routine analysis (Figure 1). The method here presented is based on a simple liquid-liquid extraction with tert-butylmethyl ether (TBME) by-passing a possible intricate solid phase extraction (SPE) procedure and a time-consuming derivatization step. The method has been fully validated according to the requirements of ISO17025/ISO15189, and its overall analytical performance has been assessed by the analysis of real samples collected from male and female subjects.

**Figure 1:**
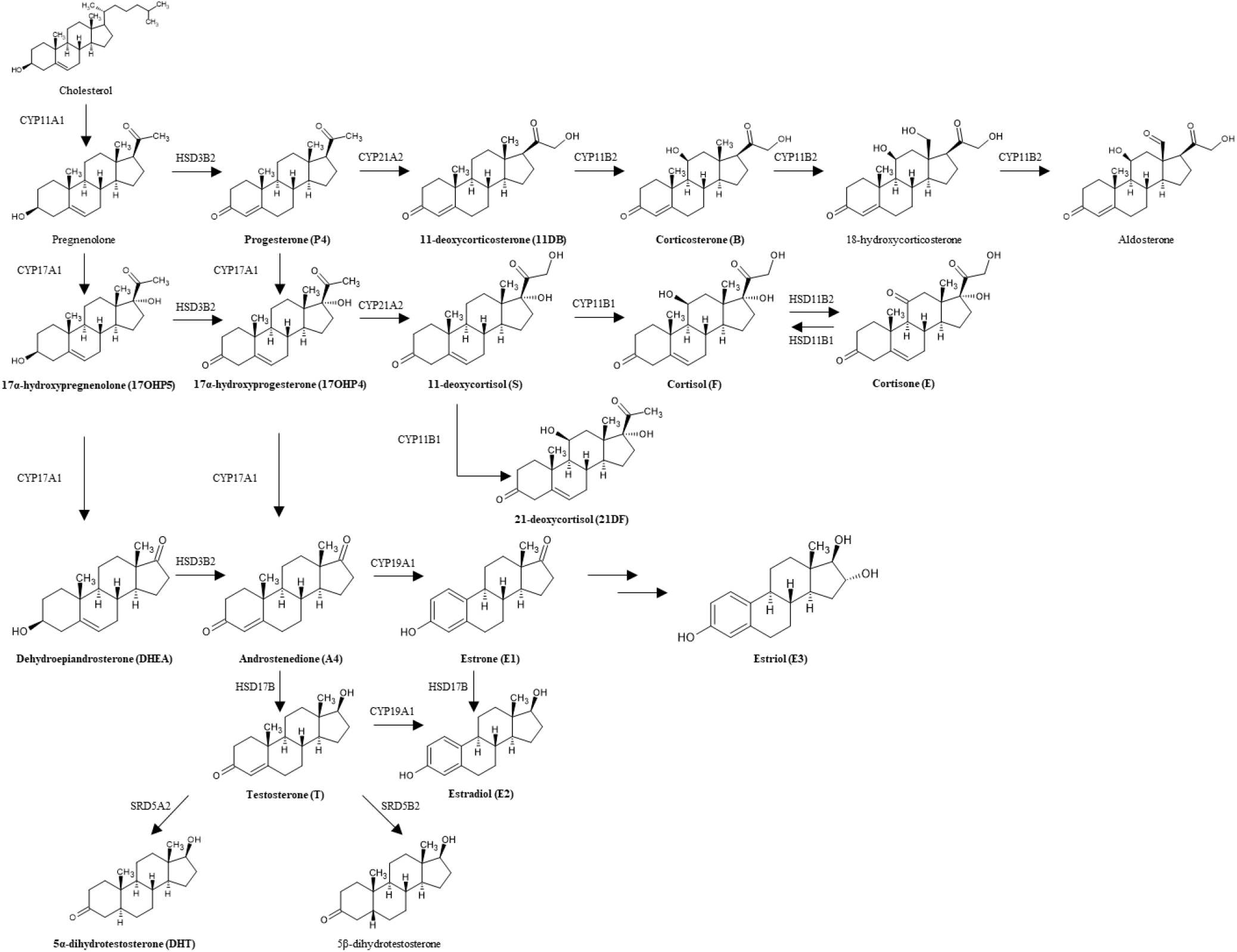
Biosynthetic pathway of endogenous steroid hormones. The name of the compounds considered in the proposed method are reported in bold. CYP11A1: P450 side-chain cleavage enzyme; HSD3B2: 3β-hydroxysteroid dehydrogenase type II; CYP21A2: 21-hydroxylase; CYP11B2: aldosterone synthase; CYP17A1: 17α-hydrolase; CYP11B1: 11β-hydroxylase; HSD11B1: 11β-hydroxysteroid dehydrogenase type I; HSD11B2: 11β-hydroxysteroid dehydrogenase type II; CYP19A1: P450 aromatase; HSD17B: 17β-hydroxysteroid dehydrogenase; SRD5A2: 5α-reductase type II; SRD5B2: 5β-reductase type II.

## Materials and methods

### Chemicals and reagents

Androstenedione (Androst-4-en-3, 17-dione; A4), dihydrotestosterone ((5α,17β)-17-Hydroxyandrostan-3-one; DHT), 17α-hydroxyprogesterone (17-Hydroxypregn-4-ene-3,20-dione; 17OHP4), 17α-hydroxypregnenolone ((3β)-3, 17-Dihydroxypregn-5-en-20-one; 17OHP5), progesterone (Pregn-4-ene-3,20-dione; P4), cortisone (17,21-Dihydroxypregn-4-ene-3, 11, 20-trione; E), cortisol ((11β)-11, 17, 21-Trihydroxypregn-4-ene-3,20-dione; F), 11-deoxycortisol (17, 21-Dihydroxypregn-4-ene-3,20-dione; S), 21-deoxycortisol ((11α)-11, 17-dihydroxypregn-4-ene-3,20-dione; 21DF), corticosterone ((11β)-11, 21-Dihydroxypregn-4-ene-3,20-dione; B), 11-deoxycorticosterone (21-Hydroxypregn-4-ene-3,20-dione; 11DB), estrone (3-Hydroxyestra-1(10),2,4-trien-17-one; E1), estradiol ((17β)-Estra-1(10),2,4-triene-3, 17-diol; E2), estriol ((16α,17β)-Estra-1,3,5(10)-triene-3, 16, 17-triol; E3) and the isotopic labeled internal standards 17α-hydroxyprogesterone-*d8* (17OHP4-*d8*), 17α-hydroxypregnenolone-*d3* (17OHP5-*d3*), cortisone-*d8* (E-*d8*), cortisol-*d4* (F-*d4*), 11-deoxycortisol-*d7* (S-*d7*), 21-deoxycortisol-*d8* (21DF-*d8*), corticosterone-*d8* (B-*d8*) and 11-deoxycorticosterone-*d7* (11DB-*d7*) were from Cambridge Isotope Laboratories Inc. (Tewksbury, MA, USA). Testosterone ((17β)-17-Hydroxyandrost-4-en-3-one; T), dehydroepiandrosterone sulfate ((3β)-17-Oxoandrost-5-en-3-yl hydrogen sulfate; DHEA-S) and the isotopic labeled internal standard progesterone-*d7* (P4-*d7*), estradiol-*d2* (E2-*d2*), testosterone-*d3* (T-*d3*) and dehydroepiandrosterone sulfate-*d5* (DHEA-S-*d5*) were purchased from Sigma Aldrich (Milano, Italy). The HPLC grade solvents (acetonitrile, methanol, tert-butylmethyl ether (TBME)), ammonium fluoride (NH_4_F), phosphate buffer (PBS) bovine serum albumin (BSA) 96% w/V and the ultra-purified water were from Sigma Aldrich (Milano, Italy). Carbonate/bicarbonate buffer solution was purchased from Carlo Erba (Milano, Italy).

### Stock solutions, calibrators and *in-house* quality control

Stock solution of each analytes and isotope labelled internal standards were prepared in methanol at concentration of 1 mg/mL. Each analyte stock solution was diluted and combined to obtain a series of working solutions in the range of concentrations 0,1 ng/mL – 100 μg/mL. The internal standard (ISTD) working solution was obtained by mixing and diluting all the isotope labelled stock solution (testosterone-*d3* (T-*d3*), dehydroepiandrosterone sulfate-*d5* (DHEA-S-*d5*), estradiol-*d2* (E2-*d2*), cortisol-*d4* (F-*d4*), cortisone-*d8* (E-*d8*), corticosterone-*d8* (B-*d8*), 11-deoxycortisol-*d7* (S-*d7*), 21-deoxycortisol-*d8* (21DF-*d8*), 11-deoxycorticosterone-*d7* (11DB-*d7*), progesterone-*d7* (P4-*d7*), 17α-hydroxyprogesterone-*d8* (17OHP4-*d8*), 17α-hydroxypregnenolone-*d3* (17OHP5-*d3*)) to obtain a final concentration of 1 ng/mL for all deuterated standards except for DHEA-S-*d5* which final concentration is of 10 ng/mL. All solutions were stored at −20°C.

Due to the lack of a steroids-free matrix, calibration samples were prepared in a surrogate matrix consisting in a solution of bovine serum albumin (BSA) in phosphate buffer solution (PBS) 4% w/V. One blank, a zero-point level and seven levels of calibrators were used for each compound (10 pg/mL – 10 ng/mL for androstenedione (A4), testosterone (T), 11-deoxycortisol (S), 21-deoxycortisol (21DF), 11-deoxycorticosterone (11DB); 40 pg/mL – 40 ng/mL for dihydrotestosterone (DHT), estrone (E1), estradiol (E2), estriol (E3), corticosterone (B), progesterone (P4), 17α-hydroxyprogesterone (17OHP4) and 17α-hydroxypregnenolone (17OHP5); 100 pg/mL – 100 ng/mL for cortisol (F) and cortisone (E); 400 pg/mL – 1000 ng/mL for dehydroepiandrosterone sulfate (DHEA-S)), according to their reference concentration ranges (Fanelli, 2011; Eisenhofer, 2017; Schiffer, 2019).

The *in-house* quality control samples (IQCs) were obtained by pooled human serum from volunteers’ donors. According to the calibration ranges considered in the present work, three levels (low, medium and high) were generated after the addition of a certain volume of methanolic working solutions to the pooled serum. For androstenedione (A4), testosterone (T), 11-deoxycortisol (S), 21-deoxycortisol (21DF), 11-deoxycorticosterone (11DB) the IQCs samples were prepared at 40 pg/mL (low), 1 ng/mL (medium) and 10 ng/mL (high); for dihydrotestosterone (DHT), estrone (E1), estradiol (E2), estriol (E3), corticosterone (B), progesterone (P4), 17α-hydroxyprogesterone (17OHP4) and 17α-hydroxypregnenolone (17OHP5) the IQCs samples were prepared at 100 pg/mL (low), 4 ng/mL (medium) and 40 ng/mL (high); for cortisol (F) and cortisone (E) the IQCs samples were prepared at 400 pg/mL (low), 10 ng/mL (medium) and 100 ng/mL (high); finally for dehydroepiandrosterone sulfate (DHEA-S) the IQCs samples were prepared at 1 ng/mL (low), 40 ng/mL (medium) and 1000 ng/mL (high). The standard working solution used to prepare the IQCs are independently with the respect to the solution used for preparation of the calibration samples.

The analysis of IQCs was necessary to assess the performances of developed method in terms of accuracy and precision.

### Sample pre-treatment

Serum samples were thawed at room temperature and together with blanks, calibration samples and IQCs freshly prepared were processed as follow: aliquots of 250 µL were transferred into 2 mL polypropylene tubes and 25 µL of ISTD working solution was added. Samples were equilibrated on a mechanical shaker for 5 min. After this period 150 µL of carbonate/bicarbonate buffer solution (pH 9,5) and 1,5 mL of TBME were added and tubes were vortexed for 5 min and the centrifuged at 10000 rpm for 10 min. After centrifugation, the organic layers were transferred into glass tubes and evaporated to dryness under nitrogen stream at 55°C. The dried extracts were reconstituted in 50 µL of water/methanol (85/15 V/V) and then injected into the LC-MS/MS system.

### Instrumentation and LC-MS/MS conditions

The liquid chromatography (LC) system consisted of an Agilent ultra-high performance liquid chromatography (UHPLC) Infinity 1290 II (Agilent Technologies, Santa Clara, USA) equipped with a G7116B autosampler, a G7120A 1290 high speed pump and coupled to an Agilent triple quadrupole tandem mass spectrometer (MS/MS) detector 6495 (Agilent Technologies, Santa Clara, USA) [https://web.uniroma1.it/dip_dms/laboratorio-lc-ms-ms]. An electrospray (ESI) ionization source, operating both in positive and in negative mode, was used. The chromatographic separation was carried out using a C18 Zorbax Eclipse Plus column (i. d: 2,1 mm; l: 50 mm; particle size: 1,8 µm, CPS Analitica, Milano, Italy). The mobile phase consisted of NH_4_F 0,2 mM in water as solvent A and of NH_4_F 0,2 mM in methanol as solvent B. The total run time was of 16 min. The gradient was set as follow: 0-2 min 15-55% B, 2-5 min 55-45% B, 5-12 min 55-70% B, 12-13 min 70-95% B. The column was flushed for 2 min at 95% B channeling the solvent flush in the waste to prevent the contamination of the capillary. Finally, the system was re-equilibrated for 3 min with 15% of B. The flow rate was set at 400 µL/min and column temperature at 40°C. An 8 sec needle wash was used between each injection.

Mass spectrometry conditions were optimized as follow: gas temperature of 200°C, capillary and noozle voltage of 3000 V and 1500 V respectively, drying gas flow of 14 L/min, sheat gas flow of 11 L/min. The nebulizer gas was set at 20 psi and the sheat gas temperature at 250°C. The ion funnel parameters were optimized as follow: high-pressure RF of 200 V (ESI +) and 90 V (ESI -), low-pressure RF of 100 V (ESI +) and 60 V (ESI -).

### Method validation

The validation of the presented analytical method was carried out according to the U. S. Food and Drug Administration (FDA) Guidelines considering the additional issues for endogenous compounds (Food and Drug Administration, 2018). The validation was accomplished in terms of selectivity/specificity, sensitivity, linearity, extraction recovery, accuracy, intra- and inter-assay precision, matrix effect, carry over and stability.

#### Selectivity and specificity

Selectivity is the extent to which the method can determine a particular compound in the analyzed matrices without interference from matrix components; specificity is defined as the ability of the method to assess, unequivocally, the analyte in the presence of other components that are expected to be present (i. e. impurities, degradation products).

To determine selectivity and specificity six blank samples, six zero calibrator samples and six samples at ± 20% of LLOQ were analyzed in the same analytical session to exclude the presence of interferences at retention times of all analytes and internal standards. These experiments were done in spiked surrogate matrices because of the endogenous nature of the steroids included in the presented method. Despite this, the ratios between quantifier/qualifiers transitions obtained after the analysis of the spiked surrogate samples were compared with the ratios between quantifier/qualifiers transitions obtained after the analysis of a methanolic standard mixture at LLOQ and of all real samples analyzed with the presented method as proof of concept.

#### Sensitivity and lower limits of quantification (LLOQs)

Sensitivity is defined as the lowest analyte concentration in the matrix that can be unambiguously identified; the lower limit of quantification (LLOQ) is the lowest amount of an analyte that can be quantitatively determined with acceptable precision and accuracy.

LLOQ was determined, for each pseudo-endogenous steroid included in the method, such as the lowest concentration with an accuracy of 80-120 %, a CV < 20 % and a five-fold response with the respect to the zero-calibrator (a blank sample to which the internal standard is added), by the analysis of at least six replicates.

#### Linearity/calibration curves

Calibration curve is the relationship between the instrument response and the calibration standards within the quantitation and linearity range of the method.

A seven-point calibration curve was analyzed for each pseudo-endogenous steroid included in the study. The linearity was determined in the range 10 pg/mL - 10 ng/mL for androstenedione (A4), testosterone (T), 11-deoxycortisol (S), 21-deoxycortisol (21DF), 11-deoxycorticosterone (11DB); in the range 40 pg/mL – 40 ng/mL for dihydrotestosterone (DHT), estrone (E1), estradiol (E2), estriol (E3), corticosterone (B), progesterone (P4), 17α-hydroxyprogesterone (17OHP4) and 17α-hydroxypregnenolone (17OHP5); in the range 100 pg/mL – 100 ng/mL for cortisol (F) and cortisone (E) and in the range 400 pg/mL – 1000 ng/mL for dehydroepiandrosterone sulfate (DHEA-S). The reported linearity ranges were investigated according to the reference concentration ranges described for each compound under study (Fanelli, 2011; Eisenhofer, 2017; Schiffer, 2019).

#### Extraction recovery

Recovery refers to the extraction efficiency of an analytical process, reported as a percentage of the known amount of an analyte carried through the sample extraction and processing step of the method. Extraction recoveries was calculated at three different levels (40 pg/ml, 1 ng/ml and 10 ng/mL for androstenedione (A4), testosterone (T), 11-deoxycortisol (S), 21-deoxycortisol (21DF), 11-deoxycorticosterone (11DB); 100 pg/mL, 4 ng/mL and 40 ng/mL for dihydrotestosterone (DHT), estrone (E1), estradiol (E2), estriol (E3), corticosterone (B), progesterone (P4), 17α-hydroxyprogesterone (17OHP4) and 17α-hydroxypregnenolone (17OHP5); 400 pg/mL, 10 ng/mL and 100 ng/mL for cortisol (F) and cortisone (E); 1 ng/mL, 40 ng/mL and 1000 ng/mL for dehydroepiandrosterone sulfate (DHEA-S)) by comparing the results obtained after the analysis of samples (n = six for each concentration levels) spiked with all the compounds considered in the study before sample pre-treatment with the results obtained after the analysis of a second aliquot of the same samples (n = six for each concentration levels) spiked with all steroid considered after liquid-liquid extraction.

#### Accuracy and precision

Accuracy is defined as the degree of closeness of the determined value to the nominal or known true value under prescribed conditions while precision is the closeness of agreement (i. e. degree of scatter) among a series of measurement obtained from multiple sampling of the same homogenous sample under the prescribed conditions.

Both accuracy and precision were investigated by the analysis of *in-house* quality control samples (IQCs, n = six for each condition) at four different concentration levels – LLOQ, low, medium and high – (10 pg/mL, 40 pg/ml, 1 ng/ml and 10 ng/mL for androstenedione (A4), testosterone (T), 11-deoxycortisol (S), 21-deoxycortisol (21DF), 11-deoxycorticosterone (11DB); 40 pg/mL, 100 pg/mL, 4 ng/mL and 40 ng/mL for dihydrotestosterone (DHT), estrone (E1), estradiol (E2), estriol (E3), corticosterone (B), progesterone (P4), 17α-hydroxyprogesterone (17OHP4) and 17α-hydroxypregnenolone (17OHP5); 100 pg/mL, 400 pg/mL, 10 ng/mL and 100 ng/mL for cortisol (F) and cortisone (E); 400 pg/mL, 1 ng/mL, 40 ng/mL and 1000 ng/mL for dehydroepiandrosterone sulfate (DHEA-S)). The analysis was performed in one day (intra-day measurement) and in three consecutive days (inter-day measurement). The values of accuracy should be 85-115 % (80-120 % for the LLOQ level) with the respect to the nominal value and precision, measured as the coefficient of variation (CV) of the measurement, should be not higher than 15 % (20 % for LLOQ level).

#### Matrix effect

The matrix effect is a direct or indirect alteration or interference in response because of the presence of unintended analytes (for analysis) or other interfering substances in the samples.

To evaluate the matrix effect, the results obtained after the analysis of six water samples fortified with all the steroids considered in the method (final concentration 1 ng/mL for androstenedione (A4), testosterone (T), 11-deoxycortisol (S), 21-deoxycortisol (21DF), 11-deoxycorticosterone (11DB); 4 ng/mL for dihydrotestosterone (DHT), estrone (E1), estradiol (E2), estriol (E3), corticosterone (B), progesterone (P4), 17α-hydroxyprogesterone (17OHP4) and 17α-hydroxypregnenolone (17OHP5); 10 ng/mL for cortisol (F) and cortisone (E) and 40 ng/mL for dehydroepiandrosterone sulfate (DHEA-S)) were compared with the results obtained after the analysis of six serum samples fortified with all the steroids considered in this work at the concentration level reported below. Results are presented in terms of percent values: a negative result indicates ion suppression while e positive result indicates ion enhancement.

#### Carry over

Carry over is defining as the appearance of an analyte in a sample from a preceding sample and was evaluated by the analysis of a blank sample and a mobile phase sample after the injection of the highest concentration level sample of the calibration curve, in three different analytical sessions.

#### Stability

Stability is defined as measure of the intactness of an analyte, described as lack of degradation in a given matrix under specific storage conditions. We evaluated the autosampler stability of the serum samples left in the autosampler of the instrument, set at 10° C, for 24 h, 48 h and 72 h.

### Proof of concept

To verify the detection capability of the developed and validated method, we analyzed serum samples collected from five males (39.8 ± 11.3) and five female volunteers (43.8 ± 12.8) and compared our results with the already published reference concentration ranges (Fanelli, 2011; Eisenhofer, 2017; Schiffer, 2019) for each steroidal marker considered in the present work.

## Results and discussion

### Method development and optimization

#### LC-MS/MS method development and optimization

To evaluate ionization source and multiple reaction monitoring (MRM) transitions, the three most intense fragments of each compound (Table 1) were selected after the analysis of pure methanolic standard solutions at concentration of 10 μg/mL both in positive and in negative ionization mode. To improve the MRM conditions, the effects of gas temperature, collision energies, fragmentor energies, drying gas flow, sheat gas flow and sheat gas temperature were also investigated. The optimal conditions are as follow: gas temperature 200°C, capillary voltage 3000 V, noozle voltage 1500 V, drying gas flow 14 L/min, sheat gas flow 11 L/min, nebulizer gas 20 psi and sheat gas temperature of 250°C.

**Table 1:** Selected *m/z* transitions for the target analytes and deuterated internal standards (ISTDs) included in the developed and validated LC-MS/MS method.

| Analyte | RT (min) | Molecular weight (uma) | Precursor ion (m/z) | Product ions (m/z) | Collision energy (eV) | Polarity |
| --- | --- | --- | --- | --- | --- | --- |
| **Target analytes** |  |  |  |  |  |  |
| E3 | 3,4 | 288,4 | 287 | 171; 145 | 40; 44 | - |
| E | 4,1 | 360,4 | 361 | 163; 91 | 24; 64 | + |
| F | 4,5 | 362,4 | 363 | 241; 121 | 40; 32 | + |
| DHEA-S | 4,7 | 368,5 | 367 | 97; 80 | 40; 40 | - |
| 21DF | 5,3 | 346,5 | 347 | 311; 105 | 20; 60 | + |
| B | 5,4 | 346,5 | 347 | 121; 329 | 40; 20 | + |
| S | 5,6 | 346,5 | 347 | 109; 97 | 40; 40 | + |
| A4 | 6,1 | 286,4 | 287 | 109; 97 | 32; 24 | + |
| E2 | 6,4 | 272,4 | 271 | 183; 145 | 52; 48 | - |
| E1 | 6,5 | 270,4 | 269 | 145; 253 | 40; 40 | - |
| 11DB | 6,8 | 330,5 | 331 | 109; 97 | 28; 40 | + |
| T | 6,9 | 288,2 | 289 | 171; 109 | 40; 32 | + |
| 17OHP4 | 7,4 | 330,5 | 331 | 109; 91 | 28; 40 | + |
| 17OHP5 | 7,5 | 330,5 | 331 | 185; 227 | 40; 28 | - |
| DHT | 8,3 | 290,2 | 291 | 255; 77 | 40; 40 | + |
| P4 | 9,3 | 314,5 | 315 | 109; 97 | 40; 20 | + |
| **ISTDs** |  |  |  |  |  |  |
| E-*d8* | 4,0 | 368,4 | 369 | 168; 124 | 28; 36 | + |
| F-*d4* | 4,3 | 366,4 | 367 | 121; 97 | 28; 32 | + |
| DHEA-S-*d5* | 4,4 | 373,2 | 372 | 98; 80 | 54; 60 | - |
| 21DF-*d8* | 5,0 | 354,5 | 355 | 319; 46 | 16; 72 | + |
| B-*d8* | 5,2 | 354,5 | 355 | 125; 337 | 24; 26 | + |
| S-*d7* | 5,5 | 353,5 | 354 | 113; 100 | 32; 40 | + |
| E2-*d2* | 6,3 | 274,2 | 273 | 184; 147 | 44; 44 | - |
| 11DB-*d7* | 6,6 | 337,4 | 338 | 113; 100 | 24; 26 | + |
| T-*d3* | 6,8 | 291,2 | 292 | 109; 97 | 44; 30 | + |
| 17OHP4-*d8* | 7,3 | 338,5 | 339 | 113; 100 | 32; 36 | + |
| 17OHP5-*d3* | 7,4 | 333,4 | 332 | 287; 109 | 20; 28 | - |
| P4-*d7* | 9,3 | 321,5 | 322 | 113; 100 | 36; 24 | + |

Different chromatographic columns were tested to study the effects of the stationary phase on the peak shapes and separation efficiency, but the best results were obtained using a 50 mm C18 column characterized by an internal diameter of 2,1 mm and a particle size of 1,8 µm. The use of this column provided a good separation selectivity also for isomeric compounds with the same precursor ion and similar product ions (i. e. corticosterone (B), 21-deoxycortisol (21DF) and 11-deoxycortisol (S)). Chromatographic separation of the endogenous steroids was then optimized by evaluating the use of several mobile phases (water, water with 0.1% of formic acid, water with NH_4_F 0,2 or 0,5 mM, acetonitrile and methanol, methanol with formic acid and methanol with NH_4_F 0,2 or 0,5 mM). The best separation was achieved using water with NH_4_F 0,2 mM as solvent A and methanol with NH_4_F 0,2 mM as solvent B for a total chromatographic run of 16 minutes. Mobile phases solvent composition plays a significant role in offering high chromatographic selectivity. For this, to increase the detection of compounds characterized by a negative ionization mode, we decided to use the NH_4_F in both aqueous and organic mobile phase (Guo 2008; Fiers, 2012).

#### Sample pre-treatment optimization

During method development and optimization, various sample pre-treatment strategies, such as protein precipitation, liquid-liquid extraction and solid-phase extraction, were considered. All the strategies are characterized by advantages and limitations. Protein precipitation with acetonitrile and formic acid (1 %) can be high-throughput but less efficient in removing phospholipids which are responsible of the matrix effect analyzing endogenous steroids is serum even if it is a crucial step during the LC-MS/MS analysis of steroid esters in blood matrices (de la Torre, 2020). Solid-phase extraction can eliminate interfering compounds from serum but requires long extraction times and could be cost-prohibitive in a high-throughput setting. Based on the above, for developing a cost-effective and high-throughput sample pre-treatment procedure characterized by repeatability and highest extraction recovery, liquid-liquid extraction was selected for the identification and quantification of the steroids included in the presented method.

Different initial volume of sample (150, 250, 500 µL), different extraction solvents (toluene, ethyl acetate and tert-buthylmethyl ether (TBME)) added in different volume (0.5, 1, 1.5, 2 mL) and different carbonate/bicarbonate buffer solution’s volume (150, 200, 250 and 500 µL) were tested. The method was finally validated using 250 µL of human serum, 150 µL of carbonate/bicarbonate buffer solution and 1.5 mL of TBME, with the latter that is the extraction solvent that provides the highest recovery.

### Method validation

The presented method was fully quantitative validated according to the U. S. Food and Drug Administration guidelines (Bioanalytical Method Validation. Guidance for Industry, 2018) in terms of selectivity/specificity, sensitivity, linearity, extraction recovery, accuracy, intra- and inter-assay precision, matrix effect, carry over and stability.

#### Selectivity and specificity

To assess the selectivity and the specificity of the presented analytical method six blank samples, six zero calibrator samples and six samples at ± 20% of LLOQ were analyzed in the same analytical session. As we can see in Figure 2 in correspondence of the retention time of all analytes, there are no peaks in the blank samples and this means that the developed method is free of interferences. Due to the endogenous origin of the analytes included in the presented method, selectivity and specificity were determined in surrogated matrices. Despite this, the ratio between quantifier ion transition and qualifier ion transitions were constantly monitored in all real samples analyzed with the presented method and compared to the calibration samples of the same batch and to the methanolic standard solutions analyzed in each session. Interference was assumed in the case of a difference greater than 30% between quantifier/qualifier ion transitions of the real sample and calibration sample.

**Figure 2:**
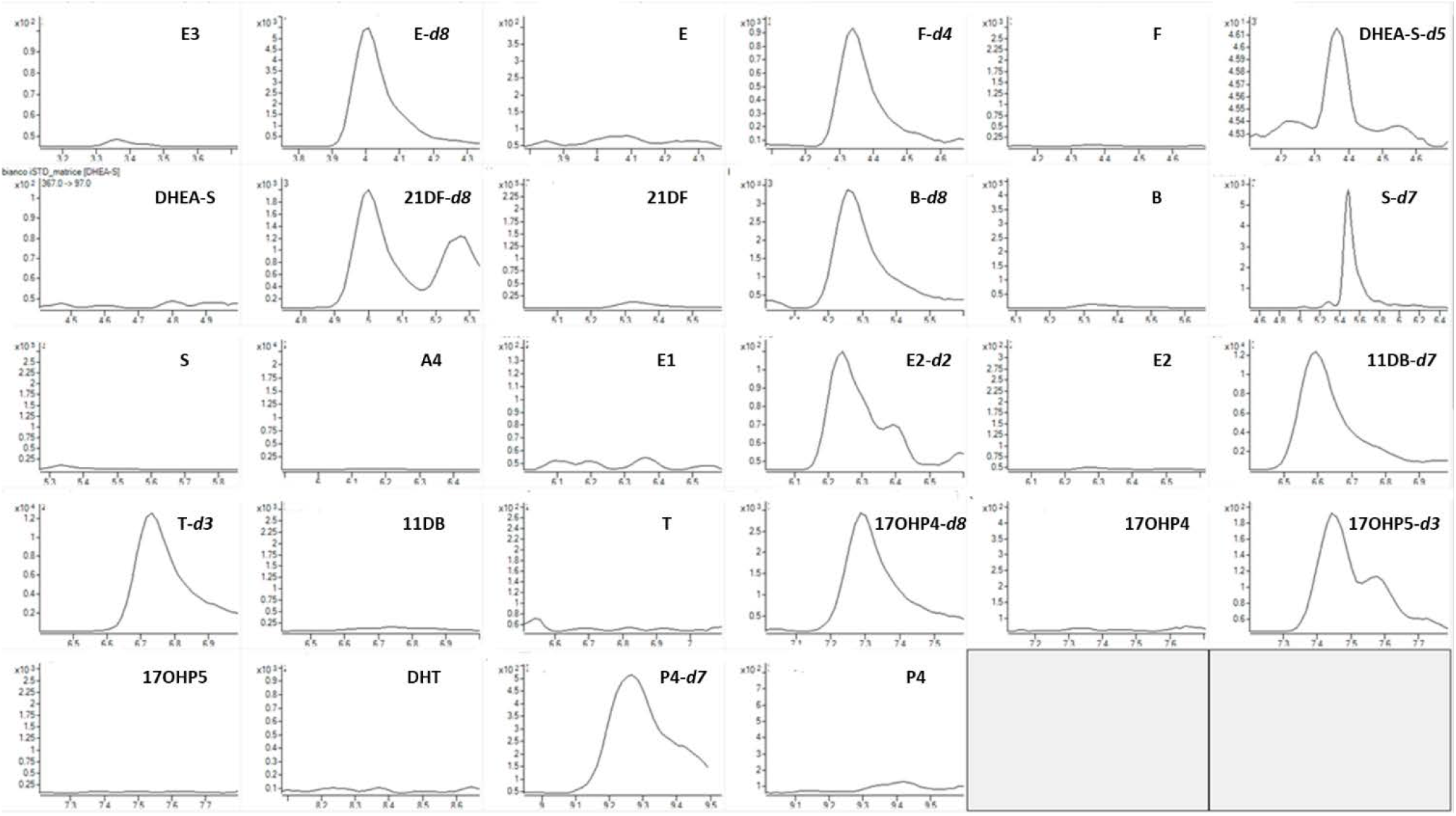
Extracted ion chromatogram of a blank serum samples fortified with all the deuterated internal standards considered in the presented analytical method (E-*d8*: 369-168, 28 eV; F-*d4*: 367-121, 28 eV; DHEA-S-*d5*: 372-98, 54 eV; 21DF-*d8*: 355-319, 16 eV; B-*d8*: 355-125, 24 eV; S-*d7*: 354-113, 32 eV; E2-*d2*: 273-185, 44 eV; 11DB-*d7*: 338-113, 24 eV; T-*d3*: 292-109, 44 eV; 17OHP4-*d8*: 339-113, 32 eV; 17OHP5-*d3*: 332-287, 20 eV; P4-*d7*: 322-113, 36 eV).

#### Sensitivity and lower limits of quantification (LLOQs)

As reported in Table 2, the LLOQs are in the range of 10 – 400 pg/mL depending on the analyte. As requested by the FDA rules, in correspondence of each LLOQ value, the accuracy of the measurement, obtained after the analysis of at least six replicates, is in the range of 80 −120 % with a CV < 20 % (Table 3). Each signal is also characterized by a five-fold response with the respect to the zero-calibrator, that is a blank sample fortified with the internal standard mixture.

**Table 2:** Lower limits of quantification (LLOQs), linearity, recovery and matrix effect for all the steroids considered in the present analytical method. The recovery concentration levels are: 40 pg/ml, 1 ng/ml and 10 ng/mL for androstenedione (A4), testosterone (T), 11-deoxycortisol (S), 21-deoxycortisol (21DF), 11-deoxycorticosterone (11DB); 100 pg/mL, 4 ng/mL and 40 ng/mL for dihydrotestosterone (DHT), estrone (E1), estradiol (E2), estriol (E3), corticosterone (B), progesterone (P4), 17α-hydroxyprogesterone (17OHP4) and 17α-hydroxypregnenolone (17OHP5); 400 pg/mL, 10 ng/mL and 100 ng/mL for cortisol (F) and cortisone (E); 1 ng/mL, 40 ng/mL and 1000 ng/mL for dehydroepiandrosterone sulfate (DHEA-S).

| Analyte | LLOQ<br>(pg/mL) | Linearity |  | Recovery %<br>(mean ± sd) |  |  | Matrix effect |
| --- | --- | --- | --- | --- | --- | --- | --- |
|  |  | Range<br>(pg/mL) | R² | Low range<br>(pg/mL) | Medium range<br>(pg/mL) | High range<br>(pg/mL) |  |
| Androgens |  |  |  |  |  |  |  |
| T | 10 | 10 - 10000 | 0,9998 | 84 ± 5,2 | 82 ± 6,3 | 88 ± 4,8 | - 7,2 % |
| DHT | 40 | 40 - 40000 | 0,9997 | 76 ± 7,7 | 78 ± 4,3 | 81 ± 5,6 | - 5,3 % |
| A4 | 10 | 10 - 10000 | 0,9994 | 73 ± 2,8 | 75 ± 4,4 | 79 ± 6,3 | - 11,8 % |
| DHEA-S | 400 | 400 - 1000000 | 0,9967 | 65 ± 4,8 | 68 ± 6,3 | 70 ± 3,5 | + 14,5 % |
| Estrogens |  |  |  |  |  |  |  |
| E1 | 40 | 40 - 40000 | 0,9997 | 70 ± 7,2 | 69 ± 4,7 | 73 ± 5,5 | - 13,2 % |
| E2 | 40 | 40 - 40000 | 0,9994 | 74 ± 2,4 | 77 ± 4,9 | 75 ± 3,4 | - 9,8 % |
| E3 | 40 | 40 - 40000 | 0,9998 | 73 ± 5,3 | 75 ± 7,6 | 79 ± 2,8 | - 14,8 % |
| Glucocorticoids |  |  |  |  |  |  |  |
| F | 100 | 100 - 100000 | 0,9997 | 72 ± 4,7 | 75 ± 6,3 | 78 ± 5,2 | + 12,4 % |
| E | 100 | 10 - 100000 | 0,9997 | 71 ± 3,1 | 72 ± 6,0 | 75 ± 4,8 | + 7,3 % |
| B | 40 | 40 – 40000 | 0,9995 | 72 ± 4,7 | 73 ± 6,4 | 79 ± 5,0 | - 8,1 % |
| S | 10 | 10 – 10000 | 0,9960 | 73 ± 4,3 | 72 ± 6,0 | 73 ± 4,9 | + 13,3 % |
| 21DF | 10 | 10 – 10000 | 0,9990 | 75 ± 3,9 | 79 ± 5,7 | 76 ± 4,4 | + 11,7 % |
| 11DB | 10 | 10 - 10000 | 0,9991 | 70 ± 3,3 | 72 ± 5,6 | 73 ± 4,9 | + 10,3 % |
| Progestagens |  |  |  |  |  |  |  |
| P4 | 40 | 40 - 40000 | 0,9991 | 78 ± 3,9 | 82 ± 6,3 | 85 ± 3,5 | - 5,3 % |
| 17OHP4 | 40 | 40 - 40000 | 0,9990 | 76 ± 3,4 | 78 ± 4,9 | 75 ± 7,5 | - 9,6 % |
| 17OHP5 | 40 | 40 - 4000 | 0,9989 | 76 ± 4,8 | 75 ± 7,5 | 76 ± 4,0 | -13,7 % |

**Table 3:** Intra- day and inter-day accuracy and precision values for all the steroid considered in the method. The values refer to four different concentration levels (LLOQ, low level, medium level and high level) characteristic of each compound.

| Analyte | Conc. | Intra-day (n = 6) % | Inter-day (n = 18) % |
| --- | --- | --- | --- |
| **Androgens** |  |  |  |
| T | LLOQ: 10 pg/mL | 85 ± 2,1 | 82 ± 3,6 |
|  | Low level: 40 pg/mL | 91 ± 4,6 | 90 ± 5,7 |
|  | Medium level: 1 ng/mL | 96 ± 5,9 | 93 ± 7,1 |
|  | High level: 10 ng/mL | 93 ± 4,5 | 92 ± 8,4 |
| DHT | LLOQ: 40 pg/mL | 82 ± 0,7 | 83 ± 0,9 |
|  | Low level: 100 pg/mL | 97 ± 10,6 | 95 ± 8,3 |
|  | Medium level: 4 ng/mL | 104 ± 9,5 | 99 ± 5,8 |
|  | High level: 40 ng/mL | 100 ± 0,7 | 98 ± 4,7 |
| A4 | LLOQ: 10 pg/mL | 89 ± 3,6 | 114 ± 6,3 |
|  | Low level: 40 pg/mL | 94 ± 4,7 | 106 ± 3,7 |
|  | Medium level: 1 ng/mL | 102 ± 8,5 | 97 ± 4,6 |
|  | High level: 10 ng/mL | 89 ± 1,5 | 87 ± 3,4 |
| DHEA-S | LLOQ: 400 pg/mL | 85 ± 3,5 | 82 ± 2,1 |
|  | Low level: 1 ng/mL | 91 ± 5,7 | 89 ± 3,6 |
|  | Medium level: 40 ng/mL | 98 ± 2,6 | 97 ± 7,2 |
|  | High level: 1000 ng/mL | 98 ± 14,3 | 96 ± 7,5 |
| **Estrogens** |  |  |  |
| E1 | LLOQ: 40 pg/mL | 94 ± 11,2 | 92 ± 8,7 |
|  | Low level: 100 pg/mL | 104 ± 11,2 | 98 ± 8,7 |
|  | Medium level: 4 ng/mL | 107 ± 7,9 | 102 ± 10,6 |
|  | High level: 40 ng/mL | 89 ± 1,2 | 87 ± 2,3 |
| E2 | LLOQ: 40 pg/mL | 99 ± 9,1 | 95 ± 6,4 |
|  | Low level: 100 pg/mL | 102 ± 5,1 | 98 ± 3,7 |
|  | Medium level: 4 ng/mL | 95 ± 3,6 | 93 ± 9,2 |
|  | High level: 40 ng/mL | 97 ± 2,9 | 93 ± 5,1 |
| E3 | LLOQ: 40 pg/mL | 85 ± 0,1 | 83 ± 1,5 |
|  | Low level: 100 pg/mL | 109 ± 7,6 | 105 ± 4,7 |
|  | Medium level: 4 ng/mL | 97 ± 6,4 | 92 ± 8,5 |
|  | High level: 40 ng/mL | 96 ± 0,5 | 94 ± 3,8 |

| Analyte | Conc. | Intra-day (n = 6) | Inter-day (n = 18) |
| --- | --- | --- | --- |
| **Glucocorticoids** |  |  |  |
| F | LLOQ: 100 pg/mL | 103 ± 1,8 | 99 ± 5,7 |
|  | Low level: 400 pg/mL | 98 ± 5,7 | 93 ± 8,6 |
|  | Medium level: 10 ng/mL | 101 ± 9,5 | 96 ± 7,2 |
|  | High level: 100 ng/mL | 95 ± 1,7 | 92 ± 5,8 |
| E | LLOQ: 100 pg/mL | 83 ± 0,5 | 82 ± 0,7 |
|  | Low level: 400 pg/mL | 92 ± 1,3 | 90 ± 3,6 |
|  | Medium level: 10 ng/mL | 98 ± 5,7 | 96 ± 2,1 |
|  | High level: 100 ng/mL | 99 ± 0,5 | 97 ± 4,6 |
| B | LLOQ: 40 pg/mL | 112 ± 3,4 | 107 ± 5,6 |
|  | Low level: 100 pg/mL | 102 ± 12,6 | 100 ± 7,8 |
|  | Medium level: 4 ng/mL | 98 ± 4,7 | 96 ± 7,3 |
|  | High level: 40 ng/mL | 104 ± 12,5 | 101 ± 3,7 |
| S | LLOQ: 10 pg/mL | 82 ± 0,3 | 81 ± 0,5 |
|  | Low level: 40 pg/mL | 99 ± 5,3 | 96 ± 1,7 |
|  | Medium level: 1 ng/mL | 102 ± 7,4 | 98 ± 8,3 |
|  | High level: 10 ng/mL | 100 ± 5,3 | 96 ± 4,6 |
| 21DF | LLOQ: 10 pg/mL | 96 ± 5,1 | 93 ± 7,8 |
|  | Low level: 40 pg/mL | 95 ± 3,7 | 92 ± 4,6 |
|  | Medium level: 1 ng/mL | 98 ± 6,2 | 100 ± 10,8 |
|  | High level: 10 ng/mL | 98 ± 3,7 | 94 ± 5,3 |
| 11DB | LLOQ: 10 pg/mL | 83 ± 1,5 | 84 ± 0,7 |
|  | Low level: 40 pg/mL | 104 ± 7,4 | 101 ± 8,5 |
|  | Medium level: 1 ng/mL | 96 ± 6,8 | 93 ± 4,7 |
|  | High level: 10 ng/mL | 99 ± 1,7 | 95 ± 5,4 |
| **Progestagens** |  |  |  |
| P4 | LLOQ: 40 pg/mL | 83 ± 1,7 | 81 ± 0,8 |
|  | Low level: 100 pg/mL | 98 ± 5,1 | 95 ± 4,2 |
|  | Medium level: 4 ng/mL | 101 ± 4,5 | 99 ± 4,6 |
|  | High level: 40 ng/mL | 94 ± 7,1 | 92 ± 8,4 |
| 17OHP4 | LLOQ: 40 pg/mL | 85 ± 1,2 | 83 ± 0,9 |
|  | Low level: 100 pg/mL | 102 ± 9,4 | 97 ± 3,6 |
|  | Medium level: 4 ng/mL | 104 ± 9,3 | 100 ± 9,7 |
|  | High level: 40 ng/mL | 100 ± 11,3 | 97 ± 7,8 |
| 17OHP5 | LLOQ: 40 pg/mL | 86 ± 3,1 | 84 ± 2,9 |
|  | Low level: 100 pg/mL | 96 ± 8,2 | 94 ± 2,4 |
|  | Medium level: 4 ng/mL | 97 ± 3,6 | 95 ± 3,9 |
|  | High level: 40 ng/mL | 99 ± 2,3 | 97 ± 7,5 |

The LLOQ values (Figure 3) are in accordance with the concentration reference ranges reported in the literature (Fanelli, 2011; Eisenhofer, 2017; Schiffer, 2019) and could be useful in the case of the analysis of serum samples collected by individual affected by metabolic disorders characterized by the reduction of concentration of endogenous steroids (i. e. hypogonadism for testosterone and other androgens or congenital adrenal hyperplasia (CAH) for cortisol and other glucocorticoids).

**Figure 3:**
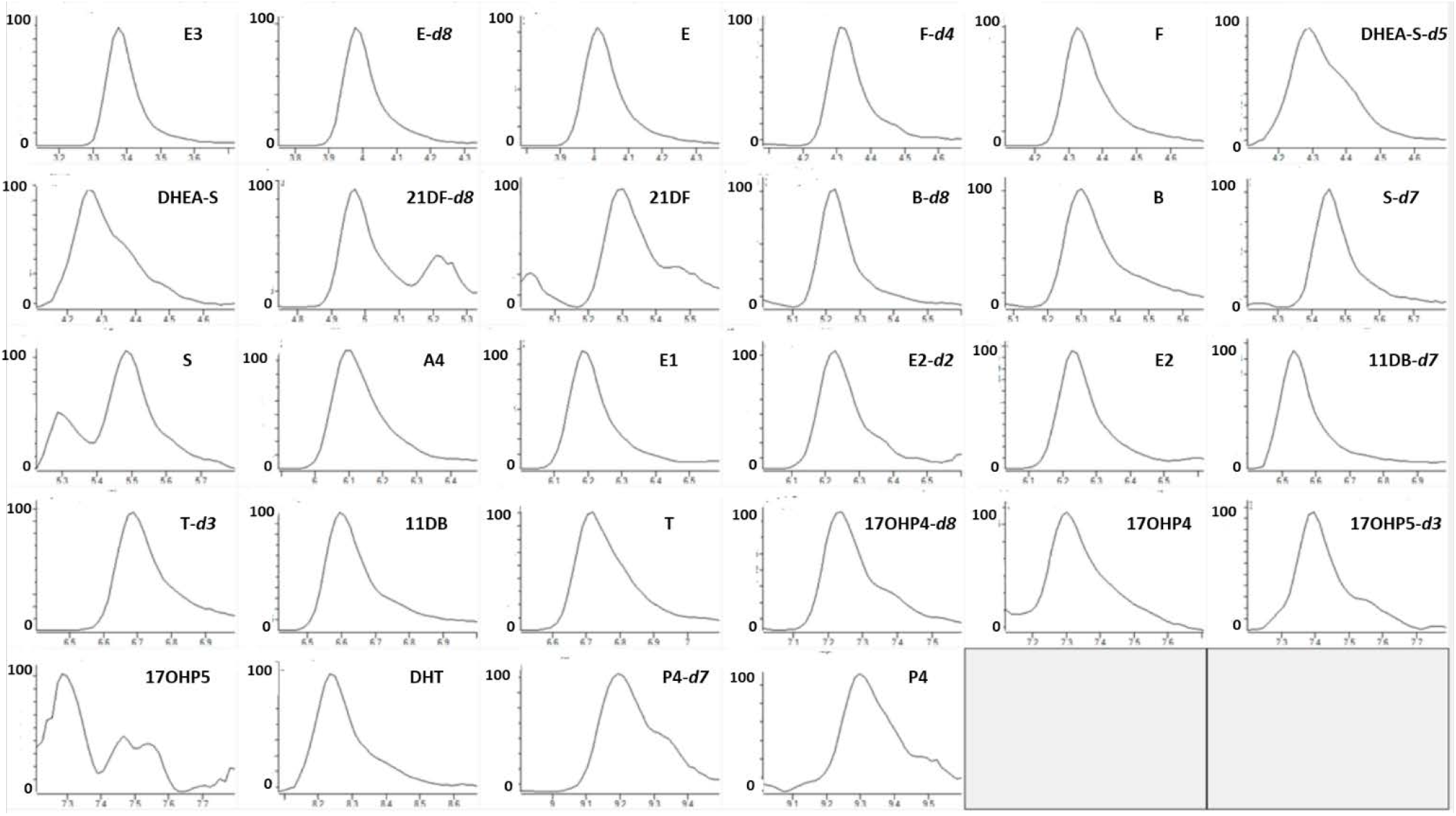
Extracted ion chromatogram of a LLOQ fortified serum samples (E3: 287-171, 40 eV; E-*d8*: 369-168, 28 eV; E: 361-163, 24 eV; F-*d4*: 367-121, 28 eV; F: 363-241, 40 eV; DHEA-S-*d5*: 372-98, 54 eV; DHEA-S: 367-97, 40 eV; 21DF-*d8*: 355-319, 16 eV; 21DF: 347-311, 20 eV; B-*d8*: 355-125, 24 eV; B: 347-121, 40 eV; S-*d7*: 354-113, 32 eV; S: 347-109, 40 eV; A4: 287-109, 32 eV; E1: 269-145, 40 eV; E2-*d2*: 273-185, 44 eV; E2: 271-183, 52 eV; 11DB-*d7*: 338-113, 24 eV; T-*d3*: 292-109, 44 eV; 11DB: 331-109, 28 eV; T: 289-171, 40 eV; 17OHP4-*d8*: 339-113, 32 eV; 17OHP4: 331-109, 28 eV; 17OHP5-*d3*: 332-287, 20 eV; 17OHP5: 331-185, 40 eV; DHT: 291-255, 40 eV; P4-*d7*: 322-113, 36 eV; P4: 315-109, 40 eV).

Furthermore, the sensitivity of the developed method, is compared to previously published assay using a higher amount of matrix (900 μL) (Fanelli, 2011) or a smaller amount of matrix (200 μL) (Peitzsch, 2015; Eisenhofer, 2017). For example, the amount on column of the steroid having the lowest LLOQ is 6 pg (Fanelli, 2015) or 0,7 pg (Peitzsch, 2015; Eisenhofer, 2017), while is 0,25 pg in the present work. This aspect underlines the good sensitivity levels obtained during the validation of the presented method.

#### Linearity/calibration curves

Due to the endogenous nature of the compounds included in the presented method, linearity experiments were assessed in surrogate matrix. To investigate the linearity, a seven points calibration’s curve was established for each steroid (10 pg/mL - 10 ng/mL for androstenedione (A4), testosterone (T), 11-deoxycortisol (S), 21-deoxycortisol (21DF), 11-deoxycorticosterone (11DB); in the range 40 pg/mL – 40 ng/mL for dihydrotestosterone (DHT), estrone (E1), estradiol (E2), estriol (E3), corticosterone (B), progesterone (P4), 17α-hydroxyprogesterone (17OHP4) and 17α-hydroxypregnenolone (17OHP5); in the range 100 pg/mL – 100 ng/mL for cortisol (F) and cortisone (E) and in the range 400 pg/mL – 1000 ng/mL for dehydroepiandrosterone sulfate (DHEA-S)) taking into account the reference concentration ranges described in the literature (Fanelli, 2011; Eisenhofer, 2017; Schiffer, 2019). In the concentration ranges here investigated all curves resulted linear with a coefficient of determination (R^2^) higher than 0,99. Furthermore, the lack-of-fit test was done for each steroid considered in the method. For each compound, the F _calc_ value was minor than F _tab_ value, confirming the linearity of the method.

#### Extraction recovery

Extraction recoveries, calculated at three different concentration levels (40 pg/ml, 1 ng/ml and 10 ng/mL for androstenedione (A4), testosterone (T), 11-deoxycortisol (S), 21-deoxycortisol (21DF), 11-deoxycorticosterone (11DB); 100 pg/mL, 4 ng/mL and 40 ng/mL for dihydrotestosterone (DHT), estrone (E1), estradiol (E2), estriol (E3), corticosterone (B), progesterone (P4), 17α-hydroxyprogesterone (17OHP4) and 17α-hydroxypregnenolone (17OHP5); 400 pg/mL, 10 ng/mL and 100 ng/mL for cortisol (F) and cortisone (E); 1 ng/mL, 40 ng/mL and 1000 ng/mL for dehydroepiandrosterone sulfate (DHEA-S) for each steroid, were between 65 and 88 % (Table 2) and then considered satisfactory for the application of the developed and validated method. The recovery was assessed as the mean (n = six replicates for each condition considered) and standard deviation of the ratio of the peaks area before and after the extraction.

#### Accuracy and precision

To evaluate both accuracy and precision of the developed method *in-house* quality control samples (IQCs, n = six for each condition) at four different concentration levels – LLOQ, low, medium and high – (10 pg/mL, 40 pg/ml, 1 ng/ml and 10 ng/mL for androstenedione (A4), testosterone (T), 11-deoxycortisol (S), 21-deoxycortisol (21DF), 11-deoxycorticosterone (11DB); 40 pg/mL, 100 pg/mL, 4 ng/mL and 40 ng/mL for dihydrotestosterone (DHT), estrone (E1), estradiol (E2), estriol (E3), corticosterone (B), progesterone (P4), 17α-hydroxyprogesterone (17OHP4) and 17α-hydroxypregnenolone (17OHP5); 100 pg/mL, 400 pg/mL, 10 ng/mL and 100 ng/mL for cortisol (F) and cortisone (E); 400 pg/mL, 1 ng/mL, 40 ng/mL and 1000 ng/mL for dehydroepiandrosterone sulfate (DHEA-S) were analyzed in three different days. The results are in accordance with the FDA criteria, underlined the applicability of the LC-MS/MS method here presented. As summarized in Table 3 the accuracy values between 85 and 115 % (80-120 % for the LLOQ level) with the respect to the nominal value and precision, measured as the coefficient of variation (CV) of the measurement, is not higher than 15 % (20 % for LLOQ level).

#### Matrix effect

Matrix effect was estimated after the analysis of a set of water samples fortified with all the steroids considered in the method at a single concentration level (1 ng/mL for androstenedione (A4), testosterone (T), 11-deoxycortisol (S), 21-deoxycortisol (21DF), 11-deoxycorticosterone (11DB); 4 ng/mL for dihydrotestosterone (DHT), estrone (E1), estradiol (E2), estriol (E3), corticosterone (B), progesterone (P4), 17α-hydroxyprogesterone (17OHP4) and 17α-hydroxypregnenolone (17OHP5); 10 ng/mL for cortisol (F) and cortisone (E) and 40 ng/mL for dehydroepiandrosterone sulfate (DHEA-S) and the analysis of a set of serum samples fortified at the same concentration. The concentration level selected is the mean point of the calibration curves analyzed in the present study. The results (Table 2) are expressed as ion enhancement (positive results) or ion suppression (negative results). The values of the matrix effect are lower than 15% for all the compounds included in the method and are adequate for the analysis of endogenous steroids in serum samples for clinical applications. These results also indicate that the optimized and selected sample pre-treatment and the use of isotopic-labelled internal do not affect the quantification of the investigated serum steroids.

#### Carry over

The evaluation of carry over was based on the analysis of a blank serum sample and of a mobile phase sample after the analysis of the highest concentration level sample of the calibration curve in three different analytical session. No steroids were detected both in the blank sample and in the mobile phase sample underlined the absence of carry over.

#### Stability

To describe the stability of our matrices, the re-analysis of a set of serum sample was performed after 24, 48 and 72 h from their pre-treatment. The samples were left in the instrument autosampler set at 10° C. The peak are ratios of each compound considered in the method were stable and reproducible and no loss of signal was registered. These results indicate that the analysis of each extract, over a reasonable period, is possible.

### Proof-of concept

Once developed and validated, the LC-MS/MS analytical method was applied to the analysis of five serum samples collected by five different male volunteers (age: 39.8 ± 11.3) and five serum samples collected by five different female healthy subjects (age: 43.8 ± 12.8). The obtained results (Table 4) are in accordance with the previously published reference ranges (Fanelli, 2011; Eisenhofer, 2017; Schiffer, 2019), demonstration the applicability of the developed procedure for the identification and quantification of serum steroids both in male and in female individuals.

**Table 4:** Serum steroid concentrations obtained after the analysis of five serum sample collected from healthy male volunteers and five serum samples collected from five healthy female volunteers. The results are expressed in term of average ± standard deviation and are compared to the reference ranges described in the literature (*: Fanelli, 2011; Eisenhofer, 2017; Schiffer, 2019).

| Analyte | Reference ranges * (ng/mL) |  | Conc. (ng/mL) in male samples (n = 5) | Conc. (ng/mL) in female samples (n = 5) |
| --- | --- | --- | --- | --- |
|  | Male | Female |  |  |
| Androgens |  |  |  |  |
| T | 0,1 - 8 | 0,1 - 3 | 2,2 ± 0,4 | 0,8 ± 0,2 |
| DHT | 0,2 - 3 | 0,05 - 2 | 0,42 ± 0,1 | 0,12 ± 0,05 |
| A4 | 0,2 - 2 | 0,1 - 3 | 0,83 ± 0,6 | 0,98 ± 0,3 |
| DHEA-S | 300 - 3000 | 300 - 3000 | 2160 ± 4,7 | 1548 ± 8,9 |
| Estrogens |  |  |  |  |
| E1 | 0,02 - 1 | 0,02 - 1 | 0,4 ± 0,01 | 0,8 ± 0,05 |
| E2 | 0,02 - 1 | 0,02 - 1 | 0,3 ± 0,05 | 0,9 ± 0,1 |
| E3 | 0,02 - 1 | 0,02 - 1 | 0,1 ± 0,04 | 0,5 ± 0,03 |
| Glucocorticoids |  |  |  |  |
| F | 45 – 250 | 40 - 250 | 140 ± 12,6 | 167 ± 9,5 |
| E | 5 - 50 | 5 - 50 | 28 ± 4,6 | 32 ± 3,3 |
| B | 0,4 - 12 | 0,4 - 25 | 3,12 ± 0,8 | 6,2 ± 0,5 |
| S | 0,04 - 1 | 0,04 – 0,5 | 0,30 ± 0,02 | 0,28 ± 0,03 |
| 21DF | < 0,1 | < 0,1 | 0,07 ± 0,01 | 0,09 ± 0,01 |
| 11DB | < 0,15 | 0,3 – 3 | 0,09 ± 0,02 | 0,42 ± 0,1 |
| Progestagens |  |  |  |  |
| P4 | 0,01 – 0,2 | 0,2 – 30 | 0,07 ± 0,02 | 2,80 ± 0,3 |
| 17OHP4 | 0,3 - 3 | 0,1 - 3 | 1,3 ± 0,2 | 1,2 ± 0,4 |
| 17OHP5 | 0,3 - 3 | 0,3 - 3 | 0,9 ± 0,2 | 1,2 ± 0,3 |

## Conclusions

Here we presented the development, validation and application of an ultrasensitive liquid chromatography – tandem mass spectrometry (LC-MS/MS) method for the simultaneous analysis of sixteen endogenous steroids in human serum. The method is specific for all compounds considered and linear in the range of concentrations investigated, that are characteristic for each analyte according to their reference population ranges. The lower limits of quantification (LLOQs) and the values of intra-day and inter-day precision and accuracy are in accordance to the FDA guidelines and no matrix effect neither carry over were observed. Finally, the presented method was applied for the analysis of serum samples collected from healthy males and females to test its application in the analysis of real samples. Based on these results, the presented method could be applied for the analysis of hormonal profile of humans subjects in whom adrenal, gonadal or combined disorders are suspected..

## Data Availability

All data are available upon reasonably motivated request.

https://isidorilab.com/home

